# AENEAS Project: Machine Vision-Based Real-Time Anatomy Detection. Application to the Pterional Trans-Sylvian Approach

**DOI:** 10.1101/2025.05.19.25327893

**Authors:** S Olei, G Sarwin, VE Staartjes, L Zanuttini, S Ryu, L Regli, E Konukoglu, C Serra

**Affiliations:** Machine Intelligence in Clinical Neuroscience (MICN) Laboratory, Department of Neurosurgery, University Hospital Zurich, Clinical Neuroscience Centre, University of Zurich, Zurich, Switzerland; Computer Vision Lab (CVL), ETH Zurich, Zurich, Switzerland; Department of Biomedical and Neuromotor Sciences (DIBINEM), University of Bologna, Bologna, Italy; Department of Neurosurgery, Daejeon Eulji University Hospital, Eulji University Medical School, Daejeon, South Korea

**Author notes:** **Correspondence:** Carlo Serra, MD, Machine Intelligence in Clinical Neuroscience (MICN) Laboratory, Department of Neurosurgery, University Hospital Zurich, Clinical Neuroscience Centre, University of Zurich, Frauenklinikstrasse 10, Zurich 8091, Switzerland. Sarwin G (equally).

## Abstract

**Introduction:** Surgical success hinges on two core factors: technical execution and cognitive planning. While the former can be trained and potentially augmented through robotics, the latter — developing an accurate “mental roadmap” of a certain operation — remains complex, deeply individualized and resistant to standardization. In neurosurgery, where minute anatomical distinctions can dictate outcomes, enhancing intraoperative guidance could reduce variability among surgeons and improve global standards. Recent developments in machine vision offer a promising avenue. Previous studies demonstrated that deep learning models could successfully identify anatomical landmarks in highly standardized procedures such as trans-sphenoidal surgery (TSS). However, the applicability of such techniques in more variable and multidimensional intracranial procedures remains unproven. This study investigates whether a deep learning model can recognize key anatomical structures during the more complex pterional trans-sylvian (PTS) approach.

**Materials and Methods:** We developed a deep learning object detection model (YOLOv7x) trained on 5.307 labeled frames from 78 surgical videos of 76 patients undergoing PTS. Surgical steps were standardized, and key anatomical targets—frontal/temporal dura, inferior frontal/superior temporal gyri, optic and olfactory nerves and internal carotid artery (ICA) — were annotated by specifically trained neurosurgical residents and verified by the operating surgeon. Bounding boxes derived from segmentation masks served as training inputs. Performance was evaluated using five-fold cross-validation.

**Results:** The model achieved promising detection performance for deep structures, particularly the optic nerve (AP_50_: 0.73) and ICA (AP_50_: 0.67). Superficial structures, like the dura and the cortical gyri, had lower precision (AP_50_ range: 0.25–0.45), likely due to morphological similarity and optical variability. Performance variability across classes reflects the complexity of the anatomical setting along with data limitations.

**Conclusion:** This study shows the feasibility of applying machine vision techniques for anatomical detection in a complex and variable neurosurgical setting. While challenges remain in detecting less distinctive structures, the high accuracy achieved for deep anatomical landmarks validates this approach. These findings mark an essential step towards a machine vision surgical guidance system. Future applications could include real-time anatomical recognition, integration with neuronavigation and the development of AI-supported “surgical roadmaps” to improve intraoperative orientation and global neurosurgical practice.

## INTRODUCTION

The success of any surgical operation depends on two factors: a cognitive factor, that is, the operator’s ability to know what should be done, according to a sort of mental “roadmap”, and a technical factor, that is, the ability to perform what the operator sets out to do. The technical factor is a consequence of the operator’s dexterity, improvable through repeated training and ideally, at least theoretically, replaceable or improvable in a future robotic context. The cognitive factor, that is, the ability to move according to a correct mental “roadmap”, on the other hand, is more complex and is a consequence of a long learning curve. This learning curve is highly individual, difficult to standardize, and is probably the main cause behind the high interindividual variability among surgeons. The same surgical procedure can be routine in the hands of one surgeon and extremely risky in the hands of another, depending on the operator’s experience (i.e. cognitive factor) with the pathology, the technique, and the surgical anatomy involved. Standardization of this cognitive process, obviously in an ameliorative sense, would allow if achieved to significantly improve the level of surgery on a global scale.

The construction of the mental roadmap recognizes several steps, of which the first is unquestionably the perfect knowledge of normal neuroanatomy and the exact understanding of pathological anatomy in general and of the individual patient in particular. It cannot be considered a coincidence that most of the technological developments introduced into the neurosurgical routine since the very beginning of this discipline (microscope, endoscope, neuronavigation, intraoperative ultrasound, electrophysiological monitoring, intraoperative magnetic resonance imaging) **(1-14)** have correct recognition of the intraoperative anatomy and discernment of normal and pathological anatomy as their ultimate goal. The recent introduction of artificial intelligence methods in medicine, pertaining particularly to the field of so-called machine vision, opens up further possibilities for improving what is already available. In a previous work we have already demonstrated how using a deep-learning-based object detection method (YOLO) applied to a large database of surgical videos related to endoscopic trans-sphenoidal pituitary surgery (TSS) it is possible to create an algorithm capable of recognizing normal anatomy visible in the nasal and sphenoidal phase of the TSS itself **(15)(16)**. The implications are obvious and promising but need further investigation and validation to fully understand its potential.

First and foremost, the question is whether the results obtained from modeling a highly repetitive and standardized surgical approach such as the TSS, where the surgical steps are repeated almost always in the same sequence, under superimposable optical conditions, with visualization of clearly distinguishable anatomical structures, are also obtainable for other neurosurgical approaches, particularly the intracranial ones.

Therefore, the purpose of this study is to evaluate (a) whether it is possible to create a machine vision algorithm that can also recognize normal intracranial brain anatomy and (b) to evaluate its performance. To this end, we chose to test this methodological approach on a database of videos of pterional trans-sylvian approaches (**PTS**), i.e., a commonly used neurosurgical procedure, whose steps are known and all in all fairly repetitive, but which presents greater variability in the sequence of these, in the morphology of the visualized anatomical structures, and above all in the optical conditions of visualization (brightness/depth/viewing angle).

## MATERIALS AND METHODS

### Data acquisition, surgical technique and labelling

In this study, we trained and validated a deep learning algorithm capable of detecting key anatomic structures encountered along a standard PTS approach. We extracted videos of PTS approach from our institutional surgical video database collecting prospectively videos of all surgeries since 2009. Surgeries were performed by one neurosurgeon (C.S.) according to the common microneurosurgical tenets. Patients head was variably rotated and tilted according to the pathology. After standard pterional craniotomy with drilling of the ala minor down to the lateral edge of the superior orbital fissure, the dura was incised and reflected anteriorly to expose sylvian fissure and related superior temporal (**STG**) and inferior frontal (**IFG**) gyri. Under the surgical microscope the basal cisterns were reached subfrontally, and the carotid as well as chiasmatic cisterns were opened thereby invariably exposing the internal carotid artery (**ICA**), the optic nerve (**II**) and the olfactory nerve (**I**) in all surgical videos.

Surgical videos frames were labelled with a dedicated software (VoTT) by specifically trained neurosurgical residents (S.O. and L.Z.) and checked according to an internal quality check protocol by the operating neurosurgeon. Both right- and left-sided cases were included. Labelling was done into 7 different classes (see also **Table 1**): superficial targets included the temporal dura mater (**TD**), the frontal dura mater (**FD**), the inferior frontal gyrus (IFG) and the superior temporal gyrus (STG). The deep targets included I, II and ICA. Given the close reciprocal proximity of some of the targets encountered in this approach, the labelling procedure was performed by coarse contour segmentation. The total amount of labels per anatomical class is shown in **Table 1**. Visual examples of labels are shown in **Figure 1**. The use of patient data from the surgical registry was approved by the local ethical review boards (KEK 2023-02265). All patients signed research consent forms.

**Table 1.**

|  | TD | FD | IFG | STG | I | II | ICA |
| --- | --- | --- | --- | --- | --- | --- | --- |
| # Labels | 1859 | 2715 | 1129 | 843 | 246 | 2307 | 1594 |

**Figure 1.**
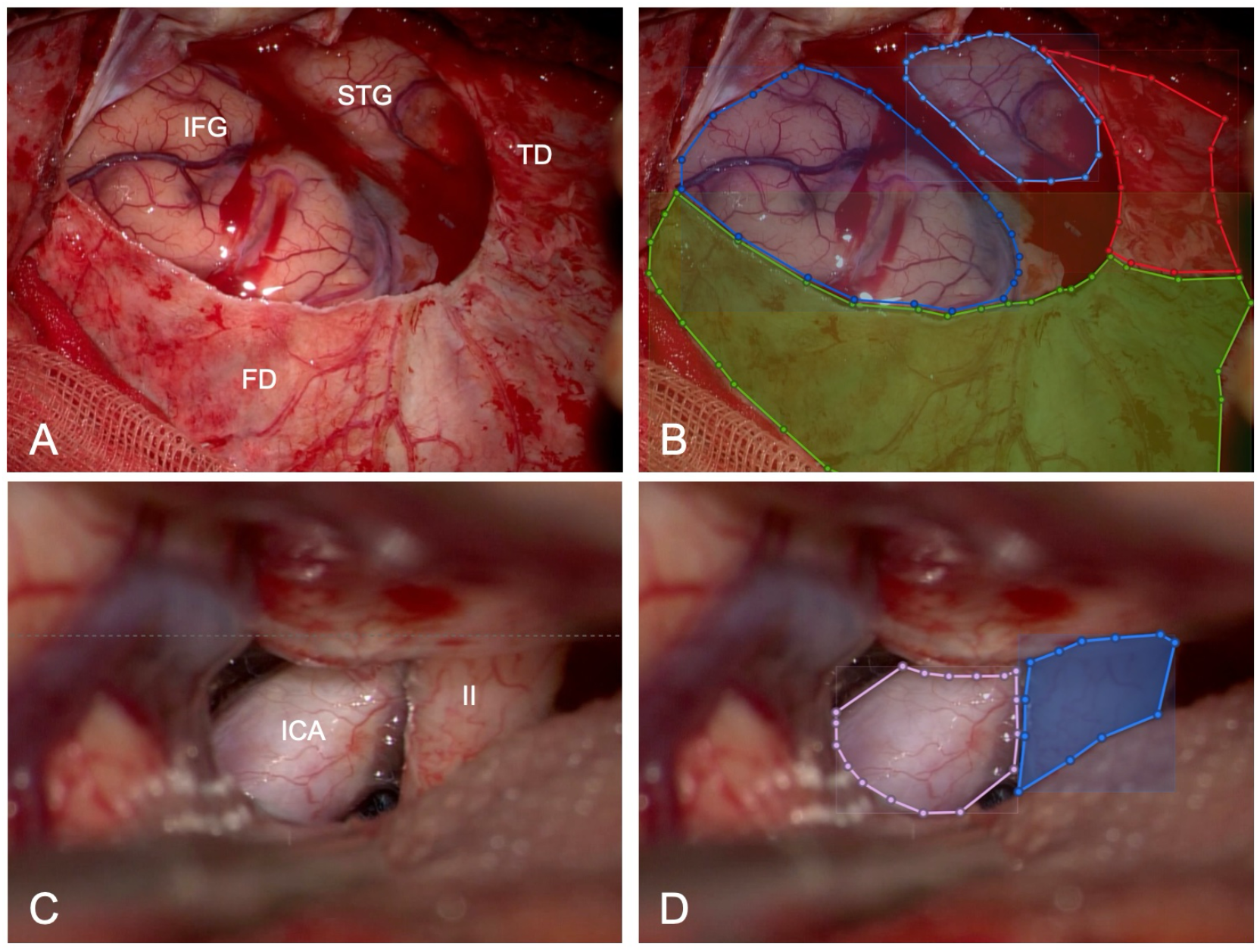
Manual labelling of anatomical targets. The anatomical segmentation and labelling of different structures is represented in this picture. Figure A shows an illustrative right-sided case, where the *frontal dura* (**FD**), *temporal dura* (**TD**), *inferior frontal gyrus* (**IFG**) and *superior temporal gyrus* (**STG**) are visualized with the surgical microscope. In figure B, the same structures are labelled with different colors using the *VoTT* software: FD is labelled with *green*, TD with *red*, IFG with *blue* and STG with *light blue*. Figure C shows an illustrative left-sided case, where the *internal carotid artery* (**ICA**) and *optic nerve* (**II**) are visualized. In figure D, the same structures are labelled: ICA with *pink* and II with *blue*.

### Model development and evaluation

As a first proof of concept, the chosen task was object detection, a problem simpler than segmentation, and therefore the segmentation masks were transformed into bounding boxes. For this task the YOLOv7x model was used. This model was trained using the standard configuration as reported in **(17)** for 300 epochs. For the evaluation of the performance the dataset was evenly, with regards to the number of patients, but randomly split into 5 different data folds. Then 5 different configurations of training, validation and test data, were used to train the network and generate object detection results. For each configuration a different fold was used as the test set, and the resulting 3 and 1 folds for training and validation, respectively. The average precision results at an intersection-over-union (IoU) threshold of 0.5 (AP_50_) for the different configurations are reported in the results section below and shown in **Table 2**.

**Table 2.**
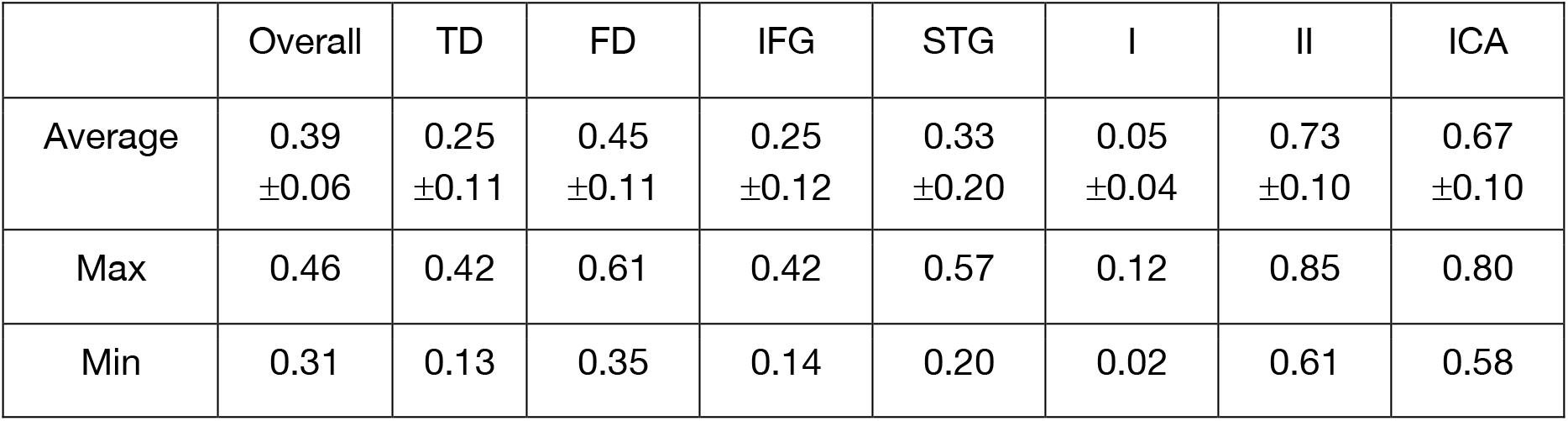

|  | Overall | TD | FD | IFG | STG | I | II | ICA |
| --- | --- | --- | --- | --- | --- | --- | --- | --- |
| Average | 0.39<br>±0.06 | 0.25<br>±0.11 | 0.45<br>±0.11 | 0.25<br>±0.12 | 0.33<br>±0.20 | 0.05<br>±0.04 | 0.73<br>±0.10 | 0.67<br>±0.10 |
| Max | 0.46 | 0.42 | 0.61 | 0.42 | 0.57 | 0.12 | 0.85 | 0.80 |
| Min | 0.31 | 0.13 | 0.35 | 0.14 | 0.20 | 0.02 | 0.61 | 0.58 |

## RESULTS

Data from a total of 78 surgical videos from 76 patients (due to re-operation in recurrent cases) undergoing PTS were collected for a total of 5.307 labelled images. Surgical indications were as follows: frontal/temporal/insular gliomas and metastases, aneurysms of the anterior circulation, craniopharyngiomas, tuberculum sellae meningiomas, tumors of the limbic system, hypothalamic tumors, meningiomas of the sphenoid wing, olfactory meningiomas, anterior crinoid meningiomas. suprasellar arachnoid cysts, third-ventricular masses, meningiomas of the planum sphenoidalis, cavernous sinus meningioma, putaminal germinoma, lymphoplasma-histioproliferative mass of the optic canal, interpeduncular chordoma, hemangiopericytoma of the middle cranial fossa, septal mass.

The achieved mean average precision (see **Table 2**) was 0.39: values were lower in case of I and in case of all superficial structures (TD, FD, IFG and STG). Interestingly however, for the remaining deep structures, the II and ICA, better AP_50_ values could be reached: 0.73 (range 0.61 – 0.85) in case of the II and 0.67 (range 0.80-0.58) in case of the ICA. Qualitative test results are visualized in **Figure 2**.

**Figure 2.**
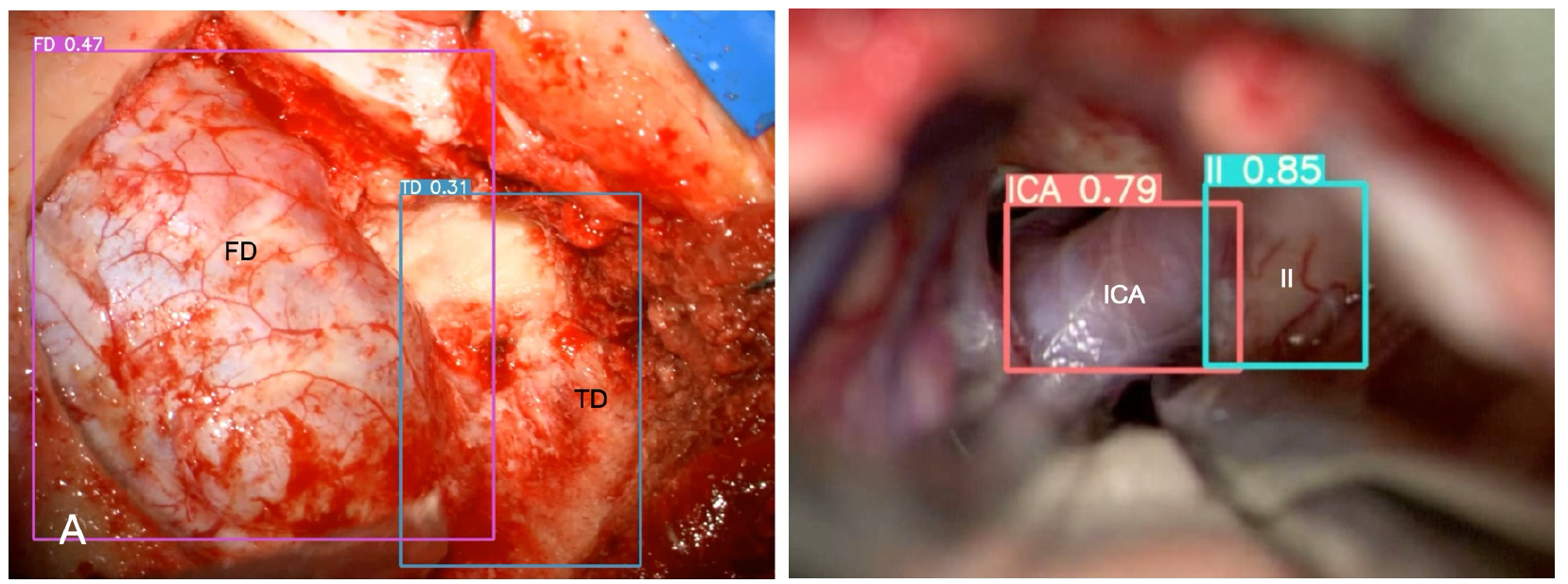
Automatic anatomical recognition. The model displays various confidence scores for superficial and deep targets. Figure A shows a right-sided illustrative case of automatic recognition through bounding boxes around *frontal* (**FD**) and *temporal dura* (**TD**), with confidence scores of 0,47 and 0,31 respectively. Figure B shows a left-sided illustrative case, where bounding boxes tightly surround the *optic nerve* (**II**) and *carotid artery* (**ICA**), displaying significantly higher confidence scores of 0,85 and 0,79 respectively. In this case, anatomical recognition is significantly more precise. The confidence score is the product of the objectness probability (how likely a box contains an object) and the highest class probability, both predicted by the model for each bounding box.

## DISCUSSION

In this preliminary study, we demonstrated how a machine vision approach can be applied to identify intracranial anatomical structures visible during a PTS approach. The novelty from a machine vision point of view is determined by the fact that the PTS approach presents, at least from a theoretical point of view, modeling difficulties that make it potentially much more difficult to analyze than the TSS endoscopic approach. However, confirmation that the same approach we have illustrated in previous publications **(15)(16)(18)** can also be used in the field of microscopic intracranial neurosurgery opens up very interesting prospects for the use of machine vision techniques, and therefore artificial intelligence in general, in improving neurosurgery by perfecting intraoperative anatomical recognition.

### Machine Vision and Neurosurgery

The use of computer vision in neurosurgery is a relatively recent development, with algorithms being developed mainly for surgical skill assessment, workflow optimization, segmenting instruments and assess the surgeon’s hand movement in the operating room **(19)(20)(21)**. Other applications of computer vision include surgical instrument tracking, with preliminary efforts addressing the real-time identification of events like brain retraction or blood loss **(22)(23)(24)**. More recent is the development of algorithms capable of identifying anatomical structures visible during endoscopic TSS, as indicated also by our research group **(15)(16)**.

However, the endoscopic TSS approach has some specific features that make it unique in its own way. The operative field is essentially a linear corridor, from the nostril to the floor of the sella through the sphenoid sinus. The anatomical structures always appear in the same sequence and almost invariably in the same three-dimensional topographical relationships with respect to the endoscope i.e., from the operator’s point of view. The endoscope itself provides a focused, high-definition visualization of the entire space being framed, and the image is transmitted in digital form to a screen for indirect viewing **(13)(14)**.

From a machine vision perspective, intracranial microneurosurgery approaches differ from trans-nasal trans-sphenoidal endoscopy in two particular aspects: firstly, the method of acquiring and viewing images is different, due to the use of a microscope instead of an endoscope. The optical characteristics of the surgical microscope impose a shallow depth of field and reduced illumination at deeper focal planes. As a result, only narrow regions are in sharp focus, while surrounding structures appear blurred or underexposed, making it difficult for models to consistently extract relevant features. Instrument occlusions in this already narrow field of view increase the complexity even further **(14)**.

Secondly, the object being viewed, i.e. the surgical field, is much more variable in terms of both the anatomical structures viewed (number of, morphological aspect etc.) and their reciprocal topographical relationships due to the greater range and freedom of movement of the microscope relative to the focal point. Microscope-assisted surgery involves frequent changes in camera angle, magnification and position, leading to inconsistent perspectives across frames. The surgeon, and therefore also the microscope, are positioned at the head of the patient, but their position, and therefore the viewing angle of the same anatomical structure, can change continuously, assuming virtually any position along the surface of the hemisphere centered on the structure under observation. To further illustrate this point, the visualized spatial relationships displayed between the internal carotid artery and the ipsilateral optic nerve or between the splenium and the Vena Galeni may radically change depending on the position of the microscope whereas such radical shifts do not occur in TSS approaches **(25)**. Therefore, the model must learn to recognize anatomical structures across a much wider range of viewpoints and spatial configurations.

### Machine Vision and Pterional Trans-sylvian Approach

In moving from a highly reproducible and controlled setting such as endoscopic TSS to the more variable setting of intracranial microneurosurgery, we chose to start with the PTS approach because, of all the intracranial approaches, it is probably the most codified and with the least extreme variability in relative positioning between the microscope and the surgical anatomy, at least in its initial steps. The aim was, in a sense, to subject our machine vision model to a kind of ‘stress test’ with more gradual changes in the key variables.

Although in this new context the machine vision system is confronted with more complex and variable conditions, the results obtained in the detection of the 7 anatomical targets considered are optimistic. In this proof-of-concept study, the application of a machine vision approach to 78 surgical videos from 76 patients, undergoing surgery through a PTS approach for different indications, allows without *a priori* anatomical knowledge to identify key anatomical targets, optic nerve and ICA, with promising accuracy. This represents a notable step forward from earlier studies from our group, showing how the same machine vision model can manage not only anatomical recognition in a relatively simple and linear surgical scenario, such as pituitary surgery, but also in a more complex tridimensional setting, such as the PTS **(26)**.

Still, AP_50_ for some more superficially located structures (TD, FD, IFG, STG) were less satisfying. This can be attributed to several factors. On one hand, the setting itself is more complex, as previously discussed. Secondly, the number of labeled images used in this experiment is approximately one-quarter of those available for the TSS approach. A third reason may be attributable to intrinsic limitations of the YOLO algorithm in clearly distinguishing certain anatomical structures from others. The fact that the algorithm has difficulty distinguishing the temporal dura from the frontal dura and the superior temporal gyrus from the inferior frontal gyrus could suggest a difficulty in recognizing structures that are visually similar from a morphological point of view and whose discrimination is based exclusively on topographical criteria. In other words, while the ICA and optic nerve are clearly distinguishable not only by their relative position in space but also by their visual appearance, the same cannot be said when differentiating between two cerebral convolutions. If the latter hypothesis were true, it would be necessary to either use other object recognition algorithms or change the input methods for the algorithm itself. Alternatively, the question arises as to whether the poor AP_50_ in structure recognition can be resolved simply by increasing the number of labeled frames. In this study, given the complex tridimensional setting in which the algorithm was applied, considering the different orientation and depth of the anatomic structures encountered throughout the operations and the small dimensions and spatial density of the deep cisternal structures, the labelling procedure on the surgical images was performed not as bounding boxes or single point annotations, but as contour segmentation. This kind of annotation procedure is time-intensive and requires attention to anatomic detail, especially when contiguous targets are considered (e.g. the optic nerve and the carotid artery), but appears to minimize inaccuracies due to similarity or possible overlapping between structures.

### Limitations and Future perspectives

The model showed promising performance in detecting the requested anatomy in the test set, particularly the optic nerve and the internal carotid artery in a common transcranial neurosurgical approach such as the PTS, representing nonetheless a complex setting due to tridimensional and operative factors. Being this study a proof-of-concept only, the number of structures tested is limited, so that they represent key landmarks for a hypothetic future „pterional roadmap” at different depths in the surgical field. An actual roadmap for such an approach would require mapping a greater number of targets, specifically in the superficial sylvian fissure splitting phase and in deeper cisternal dissection. For the same reason, only normal anatomy has been tested: recognition of pathological targets, such as tumors or aneurysms, is conceivable as a future step. Moreover, anatomic variants may further complicate the performance.

However, depending on how these issues are resolved, it is conceivable that machine vision-based algorithm could be implemented in surgical practice in the future as a tool to recognize intraoperative anatomy. It remains to be discovered, or rather investigated, whether their role will be to completely replace even the most experienced surgeons, recognizing and anticipating surgical and pathological anatomy and developing surgical strategies, or whether it will be merely complementary to current neuronavigation systems.

## CONCLUSION

With this study, we have demonstrated in a proof-of-concept setting the possibility of training machine-vision-based algorithms capable of automatically recognizing anatomical structures visible during the microsurgical trans-sylvian pterional approach. Notably, for two out of the three deeper structures (optic nerve and internal carotid artery) a very satisfactory AP_50_ score could be achieved. The results of our study also show that the accuracy of automated anatomical recognition, expressed in AP_50_, is not equally good for all anatomical structures, thus possibly highlighting the importance of the number of labels per class. Nevertheless, they represent an indispensable first step towards machine vision guided surgical roadmaps supporting intraoperative orientation in intracranial microneurosurgery.

## Data Availability

The data in the present study are not publicly accessible.

## ACKNOWLEDGEMENTS

This research is supported by SNF Project IZKSZ3_218786 research grant.

